# Is it time to ACT? A qualitative study of the acceptability and feasibility of Acceptance and Commitment Therapy for adolescents with Chronic Fatigue Syndrome

**DOI:** 10.1101/2021.04.20.21255804

**Authors:** Philippa Clery, Jennifer Starbuck, Amanda Laffan, Roxanne Parslow, Catherine Linney, Jamie Leveret, Esther Crawley

## Abstract

**Background:** Paediatric Chronic Fatigue Syndrome/Myalgic Encephalomyelitis (CFS/ME) is disabling and relatively common. Although evidenced based treatments are available, at least 15% of children remain symptomatic after one year of treatment. Acceptance and Commitment Therapy (ACT) is an alternative therapy option; however, little is known about whether it is an acceptable treatment approach. Our aim was to find out if children who are still disabled by CFS/ME after 12 months of treatment would find ACT acceptable, to inform a randomised controlled trial (RCT) of ACT.

**Methods:** We recruited children (diagnosed with CFS/ME; not recovered after one year of treatment; aged 11-17 years), their parent/carer, and healthcare professionals (HCPs) from one specialist UK paediatric CFS/ME service. We conducted semi-structured interviews to explore barriers to recovery; views on current treatments; acceptability of ACT; and feasibility of using an RCT to test effectiveness. Thematic analysis was used to identify patterns in data.

**Results:** Twelve adolescents, eleven parents, and seven HCPs were interviewed. All participants thought ACT was acceptable. Participants identified reasons why ACT might be efficacious: pragmatism, acceptance and compassion are valued in chronic illness; values-focussed treatment provides motivation and direction; psychological and physical needs are addressed; normalising difficulties is a useful life-skill. Some adolescents preferred ACT to Cognitive Behavioural Therapy as it encouraged accepting (rather than challenging) thoughts. Most adolescents would consent to an RCT of ACT but a barrier to recruitment was reluctance to randomisation. All HCPs deemed ACT feasible to deliver. Some were concerned patients might confuse ‘acceptance’ with ‘giving up’ and called for clear explanations. All participants thought the timing of ACT should be individualised.

**Conclusions:** All adolescents with CFS/ME, parents, and HCPs thought ACT was acceptable, and most adolescents were willing to try ACT. An RCT needs to solve issues around randomisation and timing of the intervention.

BOX

What is known about the subject?

1. Not all young people with CFS/ME recover.
2. ACT is a possible alternative therapy for CFS/ME, which focuses on improving functioning and quality of life rather than symptom reduction.
3. ACT is efficacious in paediatric chronic pain, and preliminary results show promising effects in adults with CFS/ME.

What this study adds?

1. ACT is an acceptable therapy for young people with CFS/ME.
2. Participants thought the ‘pragmatic’, ‘compassionate’ and ‘values-based’ focus of ACT would be helpful.
3. Adolescents, parents and healthcare professionals support a randomised controlled trial of ACT.

## Introduction

Paediatric Chronic Fatigue Syndrome/Myalgic Encephalomyelitis (CFS/ME) is relatively common (prevalence 0.55% across community, primary care and hospital populations)(1) and can be severely disabling with persistent fatigue, chronic pain, postural instability and cognitive dysfunction(2). It negatively impacts on children’s emotional(3-5), educational(6), and social functioning(7). Despite specialist treatments (Cognitive Behavioural Therapy-for-fatigue (CBT-f), Activity Management (AM) and Graded Exercise Therapy (GET)), at least 15% of children with CFS/ME remain symptomatic after one year of treatment(8). Alternative treatment approaches are needed.

Acceptance and Commitment Therapy (ACT) is an approach used in related conditions in children(9, 10). A randomised controlled trial (RCT) in paediatric chronic pain suggests ACT is better than standard care at improving functional disability and health-related quality of life(11), and recent World Health Organisation guidelines recommend ACT for treating chronic pain in children(12). Studies of ACT in CFS/ME have focussed on adults. One feasibility study in 40 adults with CFS/ME showed ACT resulted in sustained improvements in CFS/ME-related disability at six months(13).

ACT offers a similar but different approach to CBT-f(14). Differences include: focussing on improving functioning and quality of life by aligning behaviour with chosen values, rather than reducing symptoms; stepping away from thoughts (cognitive defusion) rather than challenging them; and acting presently in the moment at whatever current functional capacity is possible (psychological flexibility)(15, 16). We wanted to find out if ACT would be acceptable for children with CFS/ME.

## Methods

### Design

A qualitative study to provide multi-perspective views on ACT and a potential RCT of ACT versus treatment-as-usual.

### Recruitment

Participants were recruited from one UK specialist paediatric CFS/ME service (convenience selection). Inclusion criteria: adolescents (11-17 years) with CFS/ME(2), not recovered after one year of treatment; their parents; CFS/ME healthcare professionals (HCPs).

### Data Collection

Semi-structured interviews and one HCP focus group were undertaken (PC) February to September 2020 until data saturation was achieved(17). Participants were interviewed at home, the CFS/ME service, or over Skype. From March 2020, all were over Skype due to the COVID-19 pandemic. Adolescents and parents were asked to be interviewed separately but given the option to be together.

Topic guides were developed with psychologists (JS, AL), a qualitative researcher (RP), clinician (EC), and Young Person Advisory Group. An easy-to-understand explanation of ACT (JS, AL) was provided to participants. Questions explored: treatment needs; acceptability of ACT; and trialling ACT. HCPs were asked additional questions on delivering ACT. Interviews were checked with an experienced qualitative researcher (RP) to adapt topic guides, monitor and improve interview technique.

### Analysis

Interviews were recorded, transcribed, anonymised and imported into qualitative data-management software NVivo (PC). Notes were made during interviews. Transcripts were analysed using thematic analysis(18) to identify patterns within the data. Transcripts were double-coded (CL, AL, JS, JL) and disagreements discussed. Deductive coding was used to create a coding framework around the pre-existing ‘sensitising concepts’(19) of overarching themes “ACT acceptability” and “trialling ACT”. Inductive coding was then used to derive codes from participants’ own words to provide more detail and generate sub-themes. Data were checked between participants to explore the range of views. London-Brent Ethics Committee approved the study (19/LO/1979).

## Results

### Participants

We interviewed 30 participants: 12 adolescents (ten were female; age=12-17 years, median=15.5 years; in the service for two-five years) and 11 parents (ten were mothers; one was the parent of two adolescents). Of 14 adolescents approached, one declined to participate, one was ineligible. Three child-parent dyads were interviewed together, the remainder separately. We interviewed seven HCPs (clinicians, psychologists, physiotherapists and occupational therapists). Five took part in a focus group, two were interviewed individually. Interviews lasted 30 to 110 minutes.

### Thematic analysis

Table 2 summarises our results.

**Table 1:** Participant IDs

| ID | Child, parent or HCP |
| --- | --- |
| 01 | HCP |
| 02a | Parent |
| 02b | Child |
| 03 | HCP |
| 04a | Parent |
| 04b | Child |
| 05a | Parent |
| 05b | Child |
| 06a | Parent |
| 06b | Child |
| 07 | HCP |
| 08 | HCP |
| 09 | HCP |
| 10 | HCP |
| 11a | Parent |
| 11b | Child |
| 12a | Parent |
| 12b | Child |
| **13a** | Parent |
| **13b** | Child |
| **14** | HCP |
| **15a** | Parent |
| **15b** | Child |
| **16a** | Parent |
| **16b1** | Child |
| **16b2** | Child |
| **17a** | Parent |
| **17b** | Child |
| **18a** | Parent |
| **18b** | Child |

Key: a=parent, b=child; HCP=healthcare professional

**Table 2:** Results describing views on ACT and a potential trial presented as themes and sub-themes.

|  | **Deductive themes** | **Inductive subtheme** |
| --- | --- | --- |
| **Views on ACT** | **1) Acceptability** | An extra possibility for those struggling<br>Better than CBT-f<br>Not suitable for everyone<br>Accepting the word ‘acceptance’ |
|  | **2) Feasibility** |  |
|  | **3) Reasons why ACT could be efficacious** | Pragmatism, acceptance and compassion are valued in chronic illness<br>Cognitive defusion is less tiring but difficult to achieve<br>Focussing on values helps to “get through”<br>Addressing both psychological and physical needs<br>Normalising difficulties is a beneficial life skill |
| **Views on a trial** | **4) Barriers and facilitators to trial recruitment** | Attitudes toward research<br>Treatment fatigue<br>Reluctance to be randomised |

## 1. Acceptability

### 1.1 An extra possibility for those struggling

All 30 adolescents, parents and HCPs said ACT would “have value” (ID5a). Adolescents saw it as an “extra possibility” (ID11b) for managing CFS/ME, especially for those struggling. They felt therapy options were lacking, therefore an alternative treatment provided hope. HCPs welcomed ACT, agreeing “it’d be great to offer something else” (ID09).

> *What do we do with the kids who don’t recover? It’s a really big issue…* (ID03)

Ten of the 12 adolescents reported they would try ACT. Although, some were cautious because they were not “the biggest fan[s] of change”, they thought it was “worth trying” (ID05b) if it provided a new possibility for treatment. Two participants said they wouldn’t try ACT because they didn’t need the treatment and would be “wasting a space for someone who needs it” (ID06b) but recognised it could have been helpful for them earlier in their illness. See Supplementary Table 1 for quotes.

### 1.2 Better than CBT-f

Two participants who had already received ACT thought it was more acceptable than CBT-f because it was “more gentle and kinder” (ID13a), which was important for managing pain and fatigue. One adolescent found it “impossible” (ID11b) to challenge thoughts in CBT-f because of the cognitive effort required, so preferred the ‘values’ and ‘person-centered’ focus in ACT.

> *CBT makes you feel like you’re constantly being challenged whereas ACT just feels like it’s more accepted […]whereas CBT is trying to push you back into your old [life] despite now having a chronically ill body*. (ID13a and ID13b)

Others preferred ACT over CBT-f because it offered a “bigger picture” and “journey approach” (ID15a). One participant thought CBT-f was too focussed on “nitty-gritty” (ID15a) anxieties and could leavechildren stuck in the past. They preferred how ACT has “goal setting” and “practical elements” (ID15a) to move forward.

### 1.3 Not suitable for everyone

Parents said ACT sounded “scary” (ID06a) or “confrontational” (ID11a) for younger or timid children to dismiss thoughts (cognitive defusion), rather than explore and challenge them. In contrast, some adolescents felt this fear could be overcome: “Just the initial thought is quite scary but then after some time working on it would be okay” (ID12b). The emotional engagement required for discussing values was felt “too challenging for some people [because] talking about stuff that’s really important could upset them” (ID04b). Some questioned whether ACT was sufficiently CFS-focussed: “[ACT is for] anxiety and depression… I’d like to be explained why it would be helpful in CFS” (ID05b).

### 1.4 Accepting the word ‘acceptance’

HCPs had concerns that parents might think ACT means “you’ve just got to deal with it” (ID11a) and misunderstand ACT to be about “where you’re at now” (ID08), whereas it is “more about where you’re going, it’s still about moving things forward just through a slightly different approach.” (ID08). In their experience, parents were always searching for treatments and may find it hard to accept therapy advocating acceptance so thought the word ‘acceptance’ needed clarification.

> *It’s being really clear about what we mean by acceptance… that acceptance [is] of thoughts and commitment to that bigger life in terms of your values… but I think sometimes when people hear that word ‘acceptance’ it can feel like just putting up with things*. (ID10)

## 2. Feasibility

All HCPs felt it would be feasible to deliver ACT as it wasn’t “any more difficult” (ID08) than current psychological therapies and is currently being used, just “less formally and without a label” (ID14). However, a need for specific training was identified because “CBT is a part of core training but ACT isn’t” (ID08).

However, HCPs disagreed about *when* ACT should be offered or delivered. Some said at 12 months was not appropriate because patients may not have attended sufficient appointments by 12 months due to waiting times: “[treatment] is a year but our actual clinical contact with them is probably only six months” (ID09). They felt ACT would be more suitable for those “stuck” (ID08) after initial treatments, regardless of how long that took. Others felt ACT would be “beneficial from the get-go” (ID08) and should be offered from the beginning, not only at 12 months.

Adolescents’ opinions differed about whether ACT should be delivered *after* or *alongside* current treatments. For some, “doing the activity management and CBT [simultaneously] was too much” (ID06a), especially whilst coming to terms with the diagnosis and ‘losing’ their former life. Other adolescents reflected how their mood was inevitably affected by CFS/ME and thought psychological treatment *alongside* AM/GET would be useful. Adolescents and parents repeatedly described the importance of preventing comorbid mood disorders in CFS/ME.

> *[CFS/ME] should be looked at more holistically and [ACT] offered not just if you’re struggling with your mental health but more as a starting point*. (ID18a)

All participants agreed that the decision *if* and *when* to offer ACT should be a clinical decision “on an individual basis” (ID08) because “everyone’s different, […] what suits one person doesn’t suit another” (ID05a).

## 3. Reasons why ACT could be efficacious

### 3.1 Pragmatism, acceptance and compassion are valued in chronic illness

Participants talked about ACT being pragmatic, realistic, and accepting. They noted how thoughts and feelings around CFS/ME were valid and grounded in true events or understandable anxieties, so it was unhelpful to challenge thoughts by “changing being chronically ill to a happy thought” (ID13b). Adolescents felt compassionate acceptance was a more appropriate approach for managing the loss and grief associated with CFS/ME, than “constantly telling them to challenge feelings and distract themselves from [thoughts]” (ID13a).

### 3.2 Cognitive defusion is less tiring but difficult to achieve

Some adolescents expressed stepping away from thoughts (cognitive defusion) was a good tactic for dealing with negative cognitions and “get on with stuff” (ID15b) because constantly filtering negative thoughts exacerbated fatigue. However, some thought dismissing thoughts was too difficult. They were unsure how to subsequently deal with dismissed thoughts: “I’d be all… what… like where… what am I supposed to do with [the thought]… just leave it?” (ID12b).

### 3.3 Focusing on values helps to “get through”

Adolescents described losing “core values” (ID16b1) and thought ACT’s focus on values would be useful. They liked the practical element of committed action to values to help them “get through” their illness (ID11b).

### 3.4 Addressing both psychological and physical needs

Families felt ACT recognised the wide-ranging health and social impacts of CFS/ME. Adolescents liked ACT’s holistic “universal” (ID13b) approach to addressing both their “psychological condition, but also [ACT] helps you accept your physical one too” (ID13b).

### 3.5 Normalising difficulties is a beneficial life skill

Parents thought that ‘normalising difficulties’ in ACT was helpful to understand worries and setbacks as part of “the human condition” (ID18a) and felt that “we would all benefit from” (ID04a) these life skills. HCPs agreed that normalising difficulties is especially important for managing CFS/ME in teenagers because they “struggle with feeling weird and unique” (ID14).

See Supplementary Table 2 for illustrative quotes.

## 4. Barriers and Facilitators of trial recruitment

### 4.1 Attitude toward research

Seven of ten adolescents who said they would try ACT, said they would consent to an RCT. A key facilitator to recruitment was appreciating benefits of research. Participants expressed wanting to help others, even if the trial didn’t benefit them directly: “it’s not necessarily doing it for right now, it’s doing it for the longer-term” (ID04a). Five participants had previously participated in trials, so had insight into research involvement.

### 4.1 Treatment fatigue

Two adolescents said they wouldn’t consent to an RCT because they felt de-motivated and new treatments were “passed [them] now” (ID06b). HCPs also recognised that some might feel negative about another treatment because they “had tried everything” (ID14).

### 4.2 Reluctance to be randomised

Most understood randomisation was necessary for a trial. However, some were reluctant, stating that one RCT arm would suit them better, so if they got the opposite arm it might affect their engagement or belief in treatment efficacy. Whilst most parents also agreed to randomisation, one would prefer if their child could “have the chance to do the other [arm] afterwards […] so if [they] can [receive] both [treatments], then that would be ideal” (ID12a). Similarly, adolescents who found randomisation unacceptable said they might take part if they could subsequently receive the therapy they hadn’t received in the trial.

See Supplementary Table 3 for illustrative quotes.

## Discussion

To our knowledge, this is the first study to explore the views of adolescents with CFS/ME on ACT. All participants said ACT was acceptable and most adolescents would partake in an RCT. Parents and adolescents thought ACT was suitable for those with persistent CFS/ME symptoms because of its pragmatic and compassionate approach. Issues with delivering ACT and an RCT were discussed, including: extra training required for psychologists; timing of when ACT should be offered; and concern that patients might confuse ‘acceptance’ with ‘giving up’.

Strengths of this study include: multi-perspective views from three participant groups; interviewing adolescents with a variety of ages and illness durations; good engagement (only one adolescent declined to participate); and recruiting from the pool of adolescents who would be eligible for an RCT. Limitations are that: participants provided opinions based on information about what ACT would involve rather than actually undergoing treatment; participants were likely biased toward being engaged in treatment and research which could overestimate acceptability of ACT and the proportion who would consent to a trial; few (four) males were interviewed; and recruitment was from one UK paediatric specialist CFS/ME service, so results may not be generalisable to males or other centres.

Our findings are consistent with results from a feasibility study with children with functional somatic syndromes, where 90.5% completed group-based ACT and all would recommend it to a friend(20). In our study, some adolescents appeared to have a treatment preference for ACT or treatment-as-usual. This should be borne in mind when designing a trial.

Our study found that participants wanted pragmatic and values-focussed strategies in treatment, which is consistent with research on ACT in paediatric chronic pain(9), where the core elements of ACT (i.e., ‘functional contextualism’(21) to facilitate behaviour in line with personal values and goals(15)) have demonstrated efficacy(22). Adolescents highlighted the loss of their core-values during their illness, so perhaps values-based treatment serves as a motivational factor(23). They said a compassionate approach was also needed to address the grief and loss of sense-of-self which is common in CFS/ME(24). Similarly, they expressed the need for treatment that validates their thoughts, rather than challenges them. This is a key difference between how ACT and CBT-f approach cognitions(14, 15) and might be why some participants said they preferred ACT to CBT-f. Whilst CBT-f also enhances acceptance(25), its centrality in ACT is unique.

Comparable to adult CFS/ME literature(25-29), our study identifies ‘acceptance’ as fundamental for being able to enjoy life whilst affected by CFS/ME. Although this is common to chronic illness(30), CFS/ME presents particular challenges related to stigma, contested diagnosis and uncertain aetiology(31). In adults, it has been suggested that acceptance should be targeted before commencing other treatment, to maximise clinical benefit(26), aligning with opinions of some participants in this study who proposed ACT should be offered at the beginning of treatment.

## Conclusion

This work suggests ACT is acceptable and most adolescents and parents would consent to randomisation for a RCT. Given patients and HCPs feel there is a lack of options for those who have not yet fully recovered after receiving currently evidenced treatments, we recommend an RCT of ACT to determine its efficacy.

## Data Availability

Qualitative data available upon request (if it does not breach anonymity protocol).

## Acknowledgements

We would like to thank all those interviewed for contributing their time and thoughts.

## Funding

PC was employed on the NIHR Academic Foundation Programme at the time of research and is currently supported by the Elizabeth Blackwell Institute for Health Research, University of Bristol and the Wellcome Trust Institutional Strategic Support Fund (Grant code: 204813/Z/16/Z).

## Statement of Contribution

EC (MBChB, PhD) conceptualised the study. EC, PC (MBBS, BSc), AL (PhD), JS (PhD) and RP (PhD) contributed to study design. PC conducted data collection, data analysis and interpretation, and wrote the manuscript. CL (MA, BSc), AL, JS, RP and JL (MBBS, MSc, MSc) contributed to data analysis. All authors were involved in revisions of the manuscript and have approved it for submission.

## Competing Interests

Professor Crawley acts as a non-paid medical advisor for the Sussex and Kent ME society.

## Notes

### Author Declarations

London-Brent Ethics Committee approved the study (19/LO/1979).

