## Supplementary Tables for "Is it time to ACT? A qualitative study of the acceptability and feasibility of Acceptance and Commitment Therapy for adolescents with Chronic Fatigue Syndrome"

**Supplementary Material**

| **Table S1:** Illustrative quotes for theme 1 on participant views on acceptability of ACT and whether they would try it. | |
| --- | --- |
| **Adolescent participants who expressed they would try ACT (or had already)** | *“I think it would be interesting … like actually do it, and see what it’s like doing it.”* (ID02b)  *“Quite, like useful because it can like, because the therapy might get those thoughts and like accepting them, like you said and like being able to like do what you want even with those thoughts. […] I think it might have been helpful before I was starting back at school because I used to think that I’d never be able to go back to school.”*  (ID04b)  *“I think ACT is a good idea… I know my psychologist uses part of ACT […] I think ACT is a really good way of looking at it rather than just saying you shouldn’t do this, you shouldn’t feel like that. I think it’s a quite good approach towards it.”* (ID12b)  *“I think it has been useful […] I think it’s quite a healthy way of working […] I’ve found yeah I definitely really prefer it to the CBT.”* (ID13b)  *“I think that it would, that there’d be no harm in trying it and I feel like that it would maybe or give the opportunity for help*.” (ID15b)  *“Yeah, definitely. I think it sounds really useful. Especially for people that are like coming out of it, because there’s not really… I don’t know if there’s kind of like a therapy for like people that are like nearly out of it, like… like pretty much… like really nearly out of it. So I think it would be really, really useful, yeah. I would try it*.” (ID16b1)  *“I think [ACT] sounds like a really good idea because I imagine a lot of people who like really suffer, mentally with it. […] it sounds like quite a good idea for a lot of people to be honest.”* (ID16b2)  *“I think it sounds like a good way to help you get over things … I know that chronic fatigue does always bring up a lot of like bad thoughts and stuff. So I think that [ACT] is useful for chronic fatigue.”* (ID17b)  *“Well, it’s worth a shot, it’s definitely worth a shot – if it can help me with not only my chronic fatigue but everything else, then it’s definitely worth a shot.”* (ID18b) |
| **Adolescent participants who were nervous and cautious, but said they would try ACT** | *“It would be a very different turn around to going onto something different and I’m not the biggest fan of change…but I guess if there’s any kind of possibility of it helping me then I’m not gonna completely go no not doing it…. I think it’s worth trying to go ahead with it.”* (ID05b) |
| **Adolescent participants who said they wouldn’t try ACT at this point in their illness, but recognised its potential usefulness for others or themselves earlier on in illness** | “I wouldn’t [do ACT] now because I’m like more or less better, it would just be wasting a space for someone who needs it … but I could understand other people doing it, people who are still struggling.” (ID06b)  “I think personally if it was like a year ago or something, um, but now I feel, because I’m on medications and things I feel like it’s kind of given me a more thing of looking at things in a different perspective anyway. But if it was a year ago or something or if it was when I got ill then it definitely, yeah I definitely would have thought about it, it sounds good.” (ID11b) |

| **Table S2:** Illustrative quotes for theme 3 presenting participants’ views on why ACT might be particularly useful or efficacious for adolescents with CFS/ME . | |
| --- | --- |
| **Pragmatism, acceptance and compassion are valued in chronic illness** | “I think that compassionate element is what’s needed more than the challenging because um you know if someone’s feeling so awful that they hurt everywhere to sort of be challenged is really difficult I think and I think that’s what’s needed with the illness is compassion yeah definitely.” (ID05a)  “I feel like it would be a good way to actually accept it, what’s going on. I feel like everyone would, I think everyone would feel more relieved if they accepted it rather than just running away from it.” (ID11b)  “So like some thoughts and things are based on things, they're not just sort of random and unrealistic, they are like real worries and things […] I like that idea of accepting the thought rather than saying every like anxious thought is wrong […] ACT is a really good way of looking at it rather than just saying ‘you shouldn't do this’, ‘you shouldn't feel like that’ […] It kind of makes your feelings more valid.” (ID12b)  “yeah, it’s unnatural to um like think like try to change your opinion of being chronically ill to like a happy thought, so I think it’s definitely better” (ID13b)  *“I think it’s quite realistic, like still having chronic fatigue with you. Like you can’t just kind of forget about it. It’s always going to be there […] kind of like accepting that it’s always going to be there, it might not be present but it’s always going to have happened. I think it’s like you accept that.”* (ID16b1)  “A big thing is the loss that these young people experience and trying to challenge them on this loss with almost maybe a CBT approach actually just with the ACT it feels quite positive to .. You’re not fighting it, not necessarily fighting it, but you’re not kind of saying you’re challenging your thoughts you know it seems like you might be working together a bit and it for a non-psychologist it seems quite a positive way to go.” (ID09) |
| **Cognitive defusion is less tiring but difficult to achieve** | “I think that’s a brilliant idea about taking that away and not having to filter them I guess but just stepping back.” (ID05b)  “I think that the thoughts are still gonna be there whatever happens but I think turning away from them … still acknowledging that they're there but turning away from them and realising that you're still gonna have to get on with stuff even if they’re there. I think that’s quite a good approach to have.” (ID15b)  “if I could step away from those thoughts, I think it would be helpful, but I don’t know if I can.” (ID18b)  “I feel like it would be quite difficult to just yeah step away from it.” (ID11b)  “That does worry me slightly, like if I… if I started it like I'd be all… what… like where… what am I supposed to do with it, like how… just leave it?” (ID12b)  “Well, a lot of young people overthink very negatively so there's a positive with that” (ID14) |
| **Focusing on values helps to ‘get through’** | “I think you should definitely stay focussed on your values because that’s something that can get you through […] by having a focus, not always thinking I have to get rid of this now, but it’s your body getting through it to get to your goal”. (ID06b)  “If you’ve got something to kind of focus on it always helps me get through certain things especially and yeah having kind of values set down I feel like that would be quite helpful.” (ID11b)  “In chronic fatigue you do kind of lose some of your core values a little bit. It is quite hard to keep focused on them when you aren’t really doing anything. So I feel it’s quite good to have a reminder about like what kind of person you are, or like were, or… yeah, were before chronic fatigue. … because I feel like you probably do shift your values and what you’re focusing on quite a lot in chronic fatigue.” (ID16b1)  “Some of those people seem like or feel like they’ve lost a lot in their life then maybe that focus on values as a bigger part of therapy is maybe more helpful for those people.” (ID08)  “I think ACT sounds like it's similar to CBT but actually, perhaps there’s a little bit more of a practical element […] on being focused on values and um trying to move people um move forwards in a positive direction about looking at what motivates people. So I think, yeah, I can see it being positive **…** I think it’s a bit more like goal-setting so it gives people a focus” (ID15a) |
| **Addressing both psychological and physical needs** | **“**ACT like I guess it can help psychological conditions, but it also does help you accept your physical one too. So I think it’s like more universal” (ID13b)  “…someone telling me like how I should be doing things, and then someone more like just to talk about like how I was feeling. So it was like quite good to have like a mix of those.” (ID16b1)  “I’ve got... I’ve definitely got to do... I’ve definitely got to manage both, like, I can’t overdo it mentally and I can’t overdo it physically, I’ve just got to work things out mentally and then just... not... not limit myself, but not... not extremely on the physical side” ( ID18b) |
| **Normalising difficulties is a beneficial life skill** | “It seems like we would all benefit from it really. Everybody gets too ahead of themselves and it can take everything away. Whereas you know, to be able to sit back and think well actually “why am I thinking like this”… Any techniques to help that doesn’t involve just grabbing a tablet that makes you feel a bit calm is definitely beneficial because once you’ve learnt those techniques I imagine you can just play them throughout any situation um which is helpful, especially you know again on days when she’s frustrated because of pain and tiredness and when she wants to do it but it just feels too much. Um, you know, it’s a balance… it sounds more, like almost a bit like a life skill that is something that once you’ve– It’s not just gonna get her over this, it would keep her going. Once you’ve learnt the techniques you just carry on with those techniques. It’s not just “I’m having this because I’m ill” it’s a technique that will carry you on through” (ID04a)  “it's just important to have a full tool box so instead of just having spanner set that you have the drill set, the saw set and all of that to go with it.” (ID12a)  **“**So, I think helping him to understand that, you know, the human condition is that you have worries and things that happen that you don’t all... you know, things that you have to live with, that you have to learn.” (ID18a)  “I think normalising that idea that life will be up and down, rather than think positive all the time.. We evolve to think negatively to keep us safe – if you think you’re going to try and cross a road with positive thinking you’ll generally get knocked over. I think realism is really important.” (ID03)  “The bit that sounds really exciting for me, not just for chronic fatigue but any chronic condition, is that actually just saying well you’ve got a really you’re facing some really difficult things there’s a technique that can that’s been used to help people deal with longstanding difficult situations really, do you want to do you want to sort of try using that? If I was in that situation that would be something that would be of value that I would be up for.” (ID01)  “I think it's really important that they, that we normalise the feelings that, you know just help them understand that what they're going through, that some of the negativity that they're feeling or frustration, anger, some of the physical anger that they display…throwing phones, punching walls, is actually a normal reaction to a horrible chronic condition, you know, and the losses that they're going through. … quite a bit of work I do is around normalising… any chronic condition, any physical chronic condition can have a psychological impact.” (ID14) |

| **Table S3:** Illustrative quotes for theme 4 of participant views on barriers and facilitators of recruitment to a potential trial of ACT | |
| --- | --- |
| **Facilitators of recruitment** | |
| **Positive attitudes toward research and a willingness to help others with CFS/ME** | “I mean it’s good that you know it’s gonna in the end, it’s going to help somebody” (ID2a)  “I just want to try and help a bit with the research of it and hopefully help try and find a cure. I managed to raise about 800 pounds [for a recent CFS/ME charity event]. And I wouldn’t want someone to go through all what I have, so if we can find a cure and they didn’t have to” (ID06b)  “I think it’s worth trying to go ahead with it and just seeing. It’s just trialling it and seeing isn’t it.” (ID11a)  **“** I support any research I really do” (ID16a)  “Well, it sounds like it’s worth a try and if... if the research the helps children moving forward, then that would be great.” (ID18a)  **“**I’d like to, just to... just to see if it would work, and then it would be able to help out others […] it’s for the future of like, well, just research.” (ID18b) |
| **Participants understand the need for randomisation as part of a trial** | “If you found you were getting the [treatment trial arm] you’d already done you might be a bit like ‘oh (disappointed tone)’. But then again it’s all helping isn’t it? It’s gonna help. And if the trial comes good and the new one looks brilliant then you’re there! And if you don’t do it you’re not gonna get that far, so you know, yeah. As I say, it’s um, that’s what you’re doing when you’re taking part and that’s why you do it. Um you know it’s, like I say, if it helps, then it helps doesn’t it? That’s the aim. It’s not sort of necessarily doing it for help right now, it’s doing it for longer term, which these conditions are what they are, they do seem longer term than short.” (ID04a)  “well it’s a bit like when [name of young person] did [name of trial] because it was either GET or the pathway that she was put on in the trial” (ID05a)  “I've already done one trial and it… that, I don't think it would… there was anything that would really stop me from doing another one […] I'd be okay with [randomisation] because it's part of the study. I [think] with the [name of trial] one it was random whether you got the graded exercise or the activity management…” (ID12b)  “I mean even if it doesn’t necessarily help, like if the [trial arm] you’ve been put in isn’t the most effective, then the findings of the research would help you determine what one would actually be effective” (ID13b)  “Because like maybe one therapy wouldn’t work for one person whereas it would work really well for someone else especially if it’s randomised. I guess that’s the point of the trial” (ID16b1)  “I mean, I suppose it… you know that it’s still going to help even if you get one that… you preferred to have one of the treatments over the other one. But I suppose you’d still know that you’re helping the trial happen and you still might benefit from it.” (ID17b) |
| **Barriers to recruitment** | |
| **Treatment fatigue** | “I think because I’ve got stuff that I’m focusing on now and I’ve actually got stuff to do it’s going to make me feel better, I don’t want to speak to someone … no I don’t want to talk to anyone else. It’s passed me now.” (ID06b)  “if you’re saying you’re going to wait until they’ve not made any progress over a year, they might be pretty demotivated by then, thinking “oh God here’s another acronym, what does that stand for?” (ID03)  “…[young person] may just buckle at the thought of [another treatment or trial] because she’s tried everything and she’s been let down by her health so she’s probably quite negative about any... I think she would turn down ACT if I offered it to her.” (ID14)  “I don't know whether there's a bit of a treatment fatigue […] I think, you know, when you when you're looking at children who haven't recovered. Then, you know, sort of psychologically I'm not quite sure what place, you know, that group are in. So it might be that you have people start it and then not finish it and you know that's not very good for a research project is it.” (ID15a) |
| **Views of participants reluctant to be randomised** because they would: **(a)** want to choose which treatment arm would work better for them (ACT vs. supportive pathway), or **(b)** receive both ACT and the supportive pathway. | **(a)**  “If you’ve like already read about ACT and like you already want to do ACT and then you find out that you’re not doing it, it could like make it, the other treatment, like maybe they’ll think it’s worse and not even try to do it.” (ID04b)  “if you get one [trial arm] and you were hoping for the other [trial arm] and you feel like the other one would have worked more, that might be difficult, because obviously the other one (referring to the supportive pathway control trial arm) is like check-up every like once a year, so that would work better for me … but the other one where you’re talking to someone constantly, that wouldn’t be right for me.” (ID06b)  “I think having a choice would probably be better because you could think of what would be best for you and do that rather than just being put in it whether you like it or not.” (ID15b)  **(b)**  “I would wonder whether the [trial arm] my child was given was the right one for my child and if it isn't, whether I would have the chance to do the other [trial arm] one afterwards… Just to make sure that they get the best treatment that they possibly can… Because I… as important as research is so is my child getting better […] I think putting [patients] on the two [treatments] would make sense but obviously that doesn’t help you with your research […] I think you also need a third [arm or option], which is to combine the two [treatments] together [to see] if that’s more effective […] so if you can achieve both [treatments] then that would be my ideal” (ID12a)  “I just guess having the opportunity to switch [trial arm] if it doesn’t get better for them.” (ID11b) |
